# Comparative analysis of *Shigella* and *Campylobacter* transmission in paired longitudinal household cohort studies in urban Bangladesh and rural Tanzania

**DOI:** 10.64898/2026.09.01.26361896

**Authors:** Hannah Van Wyk, Andrew F. Brouwer, Sarah Elwood, Suporn Pholwat, Elizabeth T Rogawski McQuade, Rashidul Haque, Syed Shahnewaj Siraj Sony, Sabrina Karim Resha, Md Ohedul Islam, Mami Taniuchi, James A Platts-Mills, Joseph NS Eisenbergm

## Abstract

**Background:** Transmission pathways of enteric pathogens have been broadly characterized; however, the degree to which different pathogens exploit different transmission routes and how these patterns vary across age remain poorly understood. We quantified the relative contribution of within-household (infection sources internal to the household) and community-to-household (infection sources external to the household) transmission for *Shigella* and *Campylobacter*.

**Methods:** We conducted paired longitudinal cohort studies in urban Bangladesh and rural Tanzania, each enrolling 100 households with a child under the age of one year. Stool samples were collected from participants’ households monthly and during diarrheal episodes for one year. We estimated daily infection trajectories for each participant for *Shigella* and *Campylobacter*. We used these data in a household transmission model, estimating within-household and community-to-household transmission rates by pathogen, age group, and study site, using forward selection to identify interactions.

**Findings:** Overall, 8.8% of *Campylobacter* and 7.6% of *Shigella* infections were symptomatic. The incidence of *Shigella* and *Campylobacter* was 2.3 and 1.4 times higher, respectively, in Bangladesh compared to Tanzania. Crude associations suggest that *Campylobacter* had greater community-to-household versus within-household transmission compared to *Shigella*, but these patterns were better explained by interactions with age. Specifically, children <5 years had higher transmission rates for *Campylobacter* than *Shigella*, and, for both pathogens, higher community-to-household than within-household transmission rates. Those ≥5 years had higher *Shigella* transmission.

**Interpretation:** Apparent pathogen-specific differences in transmission patterns for *Shigella* and *Campylobacter* were driven by effect modification by age. Intervention design should consider age-specific transmission patterns along with pathogen biology.

**Funding:** The research was supported by R01 AI153254 and R01 TW012183 from the National Institutes of Health.

**Conflict of interest:** 

**Research in context:** *Evidence before this study:* Previous studies have identified person-to-person, environmental, and zoonotic transmission routes as important transmission pathways for enteric pathogens, but their relative contributions remain uncertain. On June 22, 2026, we searched PubMed using the following search terms: (“*Shigella*” OR “*Campylobacter*”) AND (“transmission route” OR “transmission path” OR “source attribution” OR “infection source”) AND (“household” OR “community” OR “environment” OR “food” OR “water” OR “person-to-person” OR “animal”) with no restrictions on language or publication date. We identified 153 records, of which 63 were considered after title and abstract screening. The vast majority of studies focused on infection source attribution for *Campylobacter* in high-income countries, emphasizing its strong environmental persistence and identifying community-acquired infections from contaminated food (typically poultry and cattle) as the primary sources of infection. By contrast, studies on *Shigella* transmission routes were limited, with most publications reporting results based on expert opinion, which typically concurred that transmission primarily occurs via person-to-person transmission and within households. The studies largely focused on biological properties of the pathogens and rarely examined how transmission patterns varied across age groups. Furthermore, no studies directly quantified or compared the relative contributions of specific transmission routes for both *Shigella* and *Campylobacter*.

*Added value of this study:* Using longitudinal household data from an urban setting (Dhaka, Bangladesh) and a rural setting (Haydom, Tanzania) combined with a mechanistic household transmission model, we quantified the relative contribution of within-household and community-to-household transmission for both *Shigella* and *Campylobacter*. In models not accounting for participant age, community-to-household transmission was more dominant for *Campylobacter* compared to *Shigella*. However, pathogen-specific differences in the relative contribution of community-to-household versus within-household did not persist after accounting for interactions between age and the other predictors. We identified substantial differences in transmission patterns across age groups, with community-to-household transmission consistently dominating among children <5.

*Implications of all the available evidence:* The literature on the transmission routes of enteric pathogens typically emphasizes pathogen biology, focusing on infectious dose or environmental persistence to understand likely transmission pathways. Known biological differences between *Campylobacter* and *Shigella* (e.g., that *Campylobacter* is more persistent in the environment and has a higher infectious dose than *Shigella*) could influence the transmission routes each pathogen can exploit, leading to assumptions that *Campylobacter* transmission is inherently driven by external sources. However, our results suggest that population-level transmission patterns cannot be inferred from pathogen biology alone. The relative importance of transmission from household members versus other sources varies substantially by age, with particularly strong contributions from non-household sources among young children. Interventions to reduce enteric pathogen transmission should consider the age-specific transmission patterns of the target population in addition to the biological characteristics of the pathogen.

## Introduction

Enteric pathogens remain a major contributor to the global burden of disease and are the third leading cause of death among children <5 years.^1^ Among these pathogens, *Shigella* and *Campylobacter* are important causes of childhood enteric infection and have been associated with intestinal inflammation and impaired linear growth in young children. However, large, comprehensive WASH trials have generally had limited effects on their transmission despite reducing some viral and protozoal infections,^2,3^ highlighting the need to better understand the transmission routes sustaining transmission of these high-burden bacterial pathogens. Transmission can occur through multiple pathways, including person-to-person, food- or waterborne, environmentally mediated, and zoonotic transmission, originating from infection sources within or outside of the household. In this study, we grouped these routes into two broader transmission components according to the location of the infection source: within-household transmission, defined as transmission (direct or indirect) attributable to an infectious household member, and community-to-household transmission, defined as transmission from an external source.

*Shigella*, due to its high infectivity and low persistence in the environment, is thought to be transmitted primarily through person-to-person transmission, consistent with a greater within-household transmission component.^4–7^ In contrast, *Campylobacter*, a zoonotic bacteria with relatively low infectivity, is thought to be primarily transmitted through point-source outbreaks arising from contamination of food, suggesting greater community-to-household transmission (e.g., originating external to the household).^8–13^ While these hypotheses about the relative contributions of within-household and community-to-household transmission for *Shigella* and *Campylobacter* are biologically plausible, population-level patterns may be driven not just by biological properties of the pathogen but also by differences in susceptibility, behavior, and exposures across hosts. In particular, heterogeneity in associations by age could produce apparent pathogen-specific differences in transmission patterns even when the relative importance of specific transmission routes is not determined by pathogen biology alone.

We applied a household transmission model to paired longitudinal cohort data from Dhaka, Bangladesh, and Haydom, Tanzania. We quantified the relative contributions of within-household and community-to-household transmission for *Shigella* and *Campylobacter* and assessed how these patterns varied by age and study site.

## Methods

### Data

Data were collected between 2022 and 2024 in the urban Mirpur neighborhood of Dhaka, Bangladesh, and the rural Haydom region of Tanzania in paired prospective longitudinal cohort studies using a shared protocol. Details of the study design have been described previously for the Tanzania cohort;^14^ the procedures relevant to the data used in this analysis were implemented identically at the Bangladesh site. Households were invited to participate in the study if they had a child under the age of one year, and enrollment continued until 100 households were recruited at each study site. Study households were visited twice weekly for one year to identify incident diarrhea in household members and monthly for routine sample collection. Diarrhea was defined as having three or more loose stools in a period of 24 hours, or one stool with visible blood. If diarrhea was identified in a household member, the frequency of collection for that household member was increased to twice weekly for the four weeks following the onset of symptoms. Stool samples were tested for the presence of *Shigella* and *Campylobacter* spp. Here, we focused our analysis on *C. jejuni*, the species responsible for the majority of enteric disease in humans.^15^ Methods used for bacterial diagnosis in the Tanzania study were described elsewhere.^14^ Briefly, participants provided approximately 5 g (solid stool) or 5 mL (liquid stool) of stool, which was collected within 6 hours of passage, transported on ice, and stored at –80 °C until total nucleic acid (TNA) extraction. TNA was extracted from a 200 mg stool aliquot using the QIAamp Fast DNA Stool Mini Kit (Qiagen, Hilden, Germany) according to the manufacturer’s instructions. Multiplex quantitative real-time polymerase chain reaction (qPCR) assays were used to detect *Campylobacter* spp. and *Shigella* using species-specific primer-probe sets.. Amplification was performed on a ViiA7 Real-Time PCR System (Thermo Fisher Scientific, Waltham, MA). The laboratory protocol used for the Bangladesh study differed in TNA extraction and multiplex qPCR procedures and is described in the Supplemental Methods.

All study participants provided informed consent for the study, and all data collection activities were approved by the institutional review boards of the University of Virginia Institutional Review Board, the National Institute for Medical Research in Tanzania, and the Ethical Review Committee and the Research Review Committee of the International Center for Diarrhoeal Disease Research, Bangladesh.

### Overview of analysis plan

Our analysis plan had two major components. First, we imputed the existing stool sample data from the study participants to generate 10,000 augmented datasets that contain the daily infection status for each study participant. These datasets were used to estimate the fraction of infections that were symptomatic, the duration of PCR positivity, and the total number of infections.

Second, we fit a household transmission model to each augmented dataset to estimate pathogen-specific community-to-household (*α*) and within-household (*β*) transmission rates adjusted for study site (preliminary model). We then used a model to estimate these transmission rates by pathogen, age group and study site (16 transmission rates total).

### PCR data and data augmentation

For each pathogen, we assumed that an individual was infectious on a given day if the PCR test for the corresponding stool sample had a cycle threshold (Ct) value of ≤30. We then used a data augmentation algorithm to impute daily infectiousness for each participant, described in the Supplemental Material. Briefly, data augmentation consisted of four steps: (1) assuming days between consecutive PCR-positive tests were also infectious, (2) stochastically sampling infection start dates and durations from constrained empirical distributions, (3) prepending a 3-day latent period to each augmented infection,^16,17^ and (4) classifying all remaining days as uninfected. To address potential misclassification based on the Ct cutoff of 30, we repeated the analysis defining infectiousness using Ct cutoffs of 35 and 27. To test the assumption that individuals were continuously infectious between consecutive positive tests, we conducted additional sensitivity analyses where the duration of PCR positivity was sampled from exponential and gamma distributions with different mean durations (20, 30, and 40 days) and allowed to vary by pathogen (see Supplemental Material).

### Mechanistic household transmission model

Our model distinguished between two sources of transmission according to whether the source was attributable to an infectious household member or not (Figure 1). We defined the community-to-household transmission rate *α*, which represents transmission from all sources external to infectious household members (e.g., person-to-person transmission from an infectious person outside of the household, externally contaminated food or water, environmental exposure, or zoonotic transmission). The within-household transmission rate *β* represents transmission routes originating from an infectious household member (e.g., person-to-person transmission from the infectious household member, environmentally mediated routes such as household fomites or food and water contaminated by the infectious household member).

**Figure 1.**
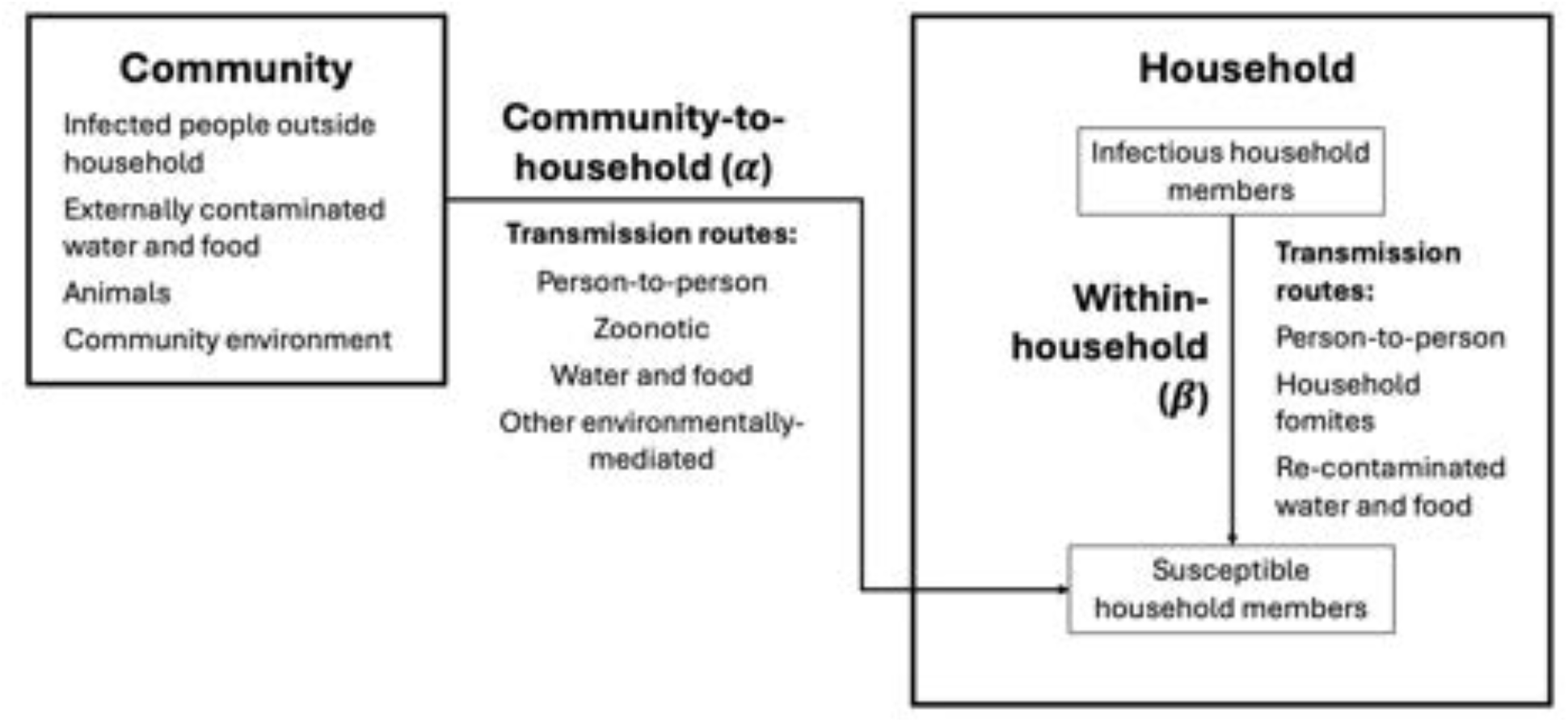
Conceptual diagram of the community-to-household and within-household transmission in the household transmission model. Community-to-household transmission (*α*) represents acquisition from all sources that are not attributable to an infectious household member. Potential sources include person-to-person transmission from infectious people outside the household, externally contaminated food or water, animals, and other community environmental reservoirs. Within-household transmission (*β*) represents infection acquisition from sources attributable to an infectious household member. Transmission from an infectious household member may similarly occur through multiple routes, including direct person-to-person transmission or environmentally mediated transmission through household fomites or food or water contaminated by the infectious household member. Thus, the two transmission rates distinguish infections according to whether acquisition is attributable to an infectious household member, rather than identifying the specific physical route through which transmission occurred.

To estimate these transmission rates, we parameterized the transmission rates on the log scale as a linear function of study site (S), pathogen (P), age group (A), transmission route (T), and potential interactions between them. Exponentiated coefficients (exp(*γ*) represent the transmission rate ratios for each covariate:

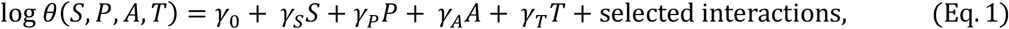

where *S, P, A*, and *T* are indicator variables for study site (*S* = 0 is Tanzania; *S* = 1 is Bangladesh), pathogen (*P* = 0 is *Shigella*; *P* = 1 is *Campylobacter*) age group (*A* = 0 is ≥5 years; *A* = 1 is <5 years), and transmission component (*T* = 0 is within-household (*β*); *T* = 1 is community-to-household (*α*)), respectively. We denote *β*_*S,P,A*_ = θ(*S, P, A*, 0) and *α*_*S,P,A*_ = θ(*S, P, A*, 1). From these terms, we define the daily force of infection (FOI), λ(*t*)_*j,ij,K*_, for each individual *i*_j_ in household *j*, for each study site (S), pathogen (P), and age group (A):

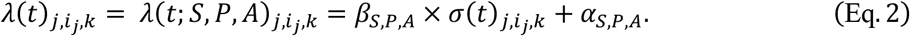

In addition to the transmission rate parameters defined in Eq. 1, the household FOI (Eq. 2) depends on *σ*(*t*)_*j,ij,K*_ which is the number of infectious people that individual *i*_*j*_ is exposed to in household *j* on day *t* in the *K*^th^ data augmentation iterate (see the Supplemental Material for more details).

To identify crude interactions between pathogen and pathway, in our preliminary model we did not adjust for age, i.e., we omitted the age group covariate and any interactions with it, but we included all pairwise interactions between pathogen, pathway, and site. In our full model, all main effects were included, and we used stepwise forward selection to select among all possible two-way interactions, iteratively adding the interaction term that produced the lowest median Bayesian Information Criterion (BIC) (see Eq. S4) relative to the equivalent model without that term. The selection process terminated when no interaction terms produced a lower median BIC. For comparison, we also estimated all parameters in a maximal model.

To estimate the parameters in the preliminary and forward-selected models, we fit a mechanistic household transmission model to each of the 10,000 augmented datasets, using the infection trajectories for each participant to calculate the likelihood of the imputed infection trajectories given the model parameters. The technical details of the household transmission model are provided in the Supplemental Material.

#### Software

Data preparation was conducted in R (v4.4; R Foundation for Statistical Computing; Vienna, Austria), and the household model likelihood was calculated in Python (v3.10.12; Python Software Foundation).

## Results

### Descriptive statistics

In Bangladesh, there were 114 enrolled households with 432 participants (mean household size: 3.75; range: 3–6) and 7407 total stool samples collected. In Tanzania, there were 100 enrolled households with 601 participants (mean household size: 5.94; range 2–12) and 7,447 stool samples collected. Four percent of stool samples were not PCR-tested from Tanzania (n=267) due to PCR equipment failure. Seven households in Bangladesh and six in Tanzania were lost to follow-up partway through the study; however, we included the available data from these households in the analysis as the households did not revoke their consent for the use of their data and the loss to follow-up should not introduce bias into the results.

Characteristics of the study participants, including the number of stools processed, results from the PCR tests, incidence of diarrhea, and sex and age distribution by study site are given in Table 1.

**Table 1.** Characteristics of the study population and percent of stool samples that were positive for *C. jejuni* and *Shigella* by age group and study site. Data were collected in Dhaka, Bangladesh, and Haydom, Tanzania, in 2022– 24.

|  | Bangladesh |  |  | Tanzania |  |  |
| --- | --- | --- | --- | --- | --- | --- |
| | Household members $\geq 5$ | Children < 5 | Total | Household members $\geq 5$ | Children < 5 | Total |
| **Number of participants** | 289 | 143 | 432 | 408 | 193 | 601 |
| **% female** | 54% | 48% | 52% | 57% | 50% | 56% |
| **Age at enrollment (yrs)** |  |  |  |  |  |  |
| Range | 5 – 75 | 0 – 4 | 0 – 75 | 5 – 77 | 0 – 4 | 0 – 77 |
| Median (IQR) | 27<br>[21, 34] | 1<br>[0, 3] | 10<br>[3, 24] | 19<br>[10, 30] | 1<br>[0, 3] | 10<br>[3, 24] |
| **Diarrhea cases, number (per person-year)** | 23<br>(0.08) | 189<br>(1.32) | 212<br>(0.49) | 7<br>(0.02) | 26<br>(0.13) | 33<br>(0.05) |
| **Stool samples, number (mean per person)** | 4023<br>(13.9) | 3384<br>(23.7) | 7407<br>(17.1) | 4688<br>(11.5) | 2492<br>(12.9) | 7180<br>(11.9) |
| **Diarrheal stools with Ct $\leq 30$ (%)** |  |  |  |  |  |  |
| *Shigella* | 23.1% | 26.8% | 26.6% | 33.3% | 33.3% | 33.3% |
| *C. jejuni* | 15.4% | 36.3% | 34.7% | 0.0% | 22.2% | 16.7% |
| **Asymptomatic stools with Ct $\leq 30$ (%)** |  |  |  |  |  |  |
| *Shigella* | 19.9% | 16.9% | 18.6% | 5.3% | 8.7% | 6.4% |
| *C. jejuni* | 13.7% | 34.8% | 22.9% | 7.9% | 20.3% | 12.2% |

### Data augmentation

Figure 2 shows data augmentation for four example households. From the 10,000 iterates of this algorithm, we used the estimated daily infection status for each study participant to obtain characteristics of the infections in the study, including: the length of PCR positivity, the incidence rates, and the symptomatic percentages (Table 2). The empirically estimated length of PCR positivity had a median of 32 days (IQR [19, 48]) for *C. jejuni* and 29 days (IQR [17, 44]) for *Shigella*. The figures showing the density of the estimated infection lengths for each pathogen can be found in Figure S3.

**Table 2.** Incidence rate (number of infections per person-year) and percent of infections that were symptomatic with 95% credible intervals for each pathogen across the two countries, stratified by age group.

|  |  |  | Household members<br>≥5 | Children < 5 | Overall |
| --- | --- | --- | --- | --- | --- |
| **Incidence** | Bangladesh | *C. jejuni* | 1.27 (1.26, 1.28) | 2.46 (2.45, 2.47) | 1.66 (1.66, 1.67) |
|  |  | *Shigella* | 1.74 (1.73, 1.75) | 1.77 (1.76, 1.78) | 1.74 (1.74, 1.75) |
|  | Tanzania | *C. jejuni* | 0.58 (0.56, 0.59) | 1.95 (1.90, 1.99) | 1.20 (1.18, 1.22) |
|  |  | *Shigella* | 0.65 (0.63, 0.66) | 1.01 (0.99, 1.02) | 0.77 (0.75, 0.78) |
| **Symptomatic (%)** | Bangladesh | *C. jejuni* | 1.7% (1.7, 1.8) | 24.7% (24.5, 24.9) | 13.3% (13.3, 13.4) |
|  |  | *Shigella* | 2.2% (2.2, 2.2) | 21.8% (21.7, 22.1) | 9.7% (9.7, 9.8) |
|  | Tanzania | *C. jejuni* | 1.3% (1.3, 1.3) | 6.5% (5.7, 7.0) | 4.1% (3.7, 4.4) |
|  |  | *Shigella* | 2.2% (2.1, 2.2) | 6.9% (6.8, 7.1) | 4.3% (4.2, 4.3) |

**Figure 2.**
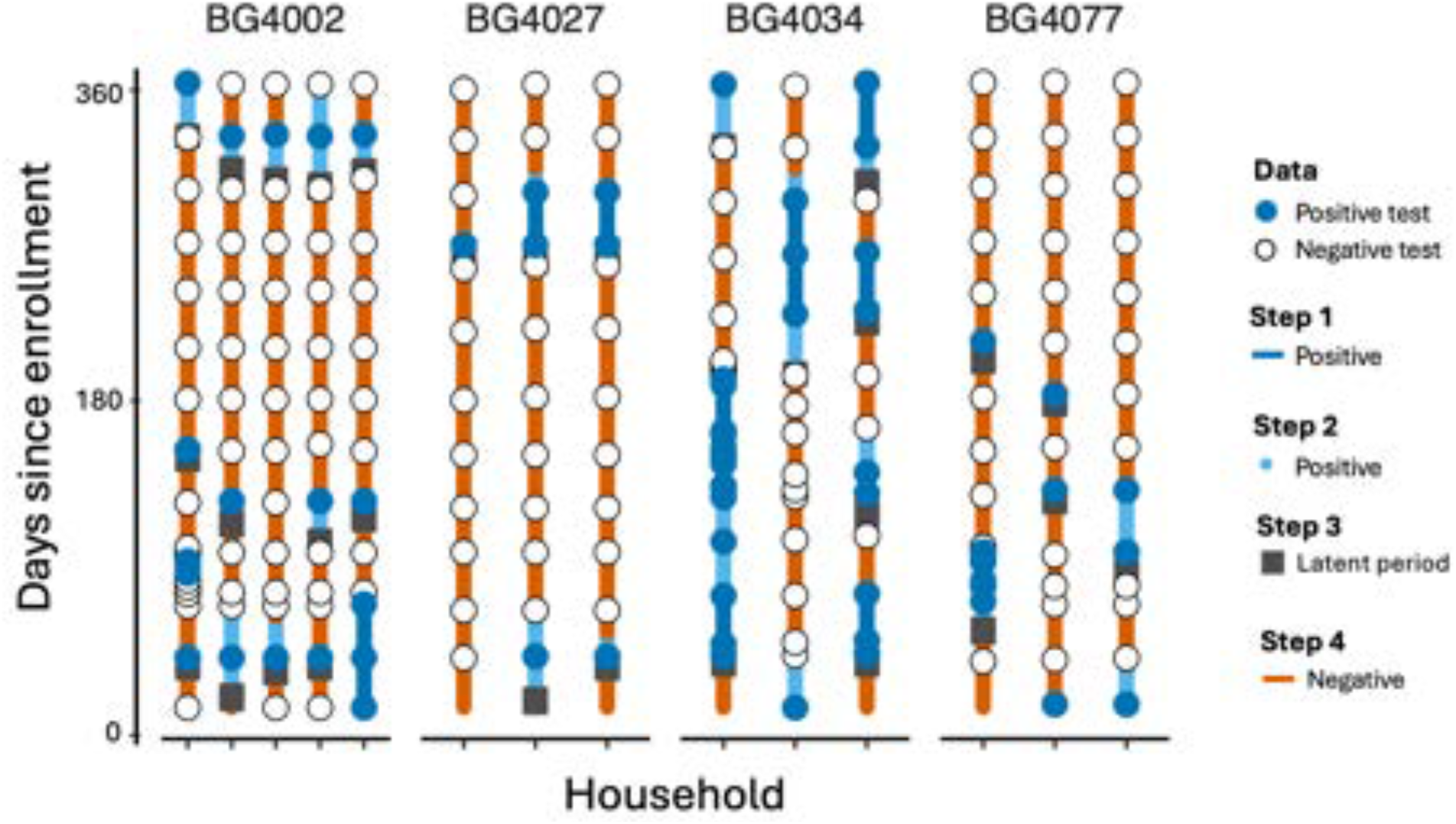
Data augmentation example for *Shigella* in four households in Bangladesh. Each line represents a household member over the course of enrollment. Circles correspond to PCR test data: white circles are negative tests (Ct > 30) and blue circles are positive tests (Ct ≤ 30). Data were augmented with a four-step process. **Step 1:** days were inferred to be infectious because they were between two consecutive positive tests (dark blue lines). **Step 2:** infections (dark blue lines) were augmented by pulling the true infection length from an empirical infection length distribution (see Figure S3) (light blue line segments). **Step 3:** three days of latency were prepended to each infection (green segments). **Step 4:** remaining days were assumed to be uninfected (orange lines). Increased test frequency is due to diarrheal episodes; otherwise tests were collected monthly.

Incidence rates for both pathogens were higher in Bangladesh. Children aged <5 years had a higher incidence of *Campylobacter* in both countries, while household members ≥ 5 years had a higher incidence of *Shigella* (compared to *Campylobacter*). We calculated low levels of symptomatic infections (across both sites and age groups, only 8.8% of *Campylobacter* and 7.6% of *Shigella* infections were symptomatic), though a greater fraction of infections with either pathogen were symptomatic among younger children in Bangladesh (20–25%) compared to older children and adults in either site (1–2%). In both Tanzania and Bangladesh, children <5 years had higher proportions of symptomatic infections than household members ages ≥5 years.

### Household transmission model

#### Preliminary model

We first estimated the pathogen–pathway interaction without adjusting for confounding or effect modification by age group; (see Figure S4, Tables S1 and S2). The ratio of the community-to-household to within-household transmission rates was 2.05-fold higher for *C. jejuni* compared to *Shigella* (pathogen-by-pathway rate ratio: 2.05 [95% CI: 1.69, 2.49]). In this model, the ratio of community-to-household to within-household transmission rates for *Shigella* was 1.53 [95% CI 1.35, 1.74] and 0.73 [95% CI: 0.60, 0.84] in Bangladesh and Tanzania, respectively, while these ratios for *Campylobacter* were 3.15 [95% CI 2.66, 3.78] in Bangladesh and 1.51 [95% CI: 1.29, 1.77] in Tanzania (each 2.05 times those of *Shigella)*.

#### Forward-selected final model

All pairwise interaction terms *except* for the pathogen–pathway interaction were selected into the final model. This model had 16 transmission rate parameters (two transmission pathways stratified by four factors) ranging from 0.86 (95% CrI: 0.79, 0.95) infections per 1,000 susceptible people per day (for *Campylobacter* community-to-household transmission rate in Tanzanian household members ages ≥5) to 8.31 (95% CrI: 8.07, 8.53) (for *Campylobacter* community-to-household transmission rate in Bangladeshi children <5). The model rate ratios and values of the 16 transmission rate parameters can be found in Supplemental Tables S2 and S3, respectively.

The median values of the 16 transmission rates are summarized in Figure 3 as a series of paired lollipop plots, with each panel connecting analogous median estimates that differed by study site (Figure 3A), pathogen (Figure 3B), transmission pathway (Figure 3C), or age group (Figure 3D). Consistent with the higher incidence of both *Shigella* and *Campylobacter* in Bangladesh (Table 2), each subgroup of age, pathogen, and transmission pathway had a higher transmission rate in Bangladesh than Tanzania (Figure 3A). Children <5 always had higher transmission for *Campylobacter* than *Shigella*, while *Shigella* transmission dominated for those ages ≥5 (Figure 3B). We also found that the community-to-household transmission rate was always higher than the within-household transmission rate for children <5. For those ages ≥5 in Tanzania, the within-household transmission rates were slightly higher than the community-to-household transmission rates (Figure 3C). When comparing across age groups, children <5 generally had higher transmission rates, except for within-household *Shigella* transmission, where household members ≥5 years had higher transmission rates (Figure 3D).

**Figure 3.**
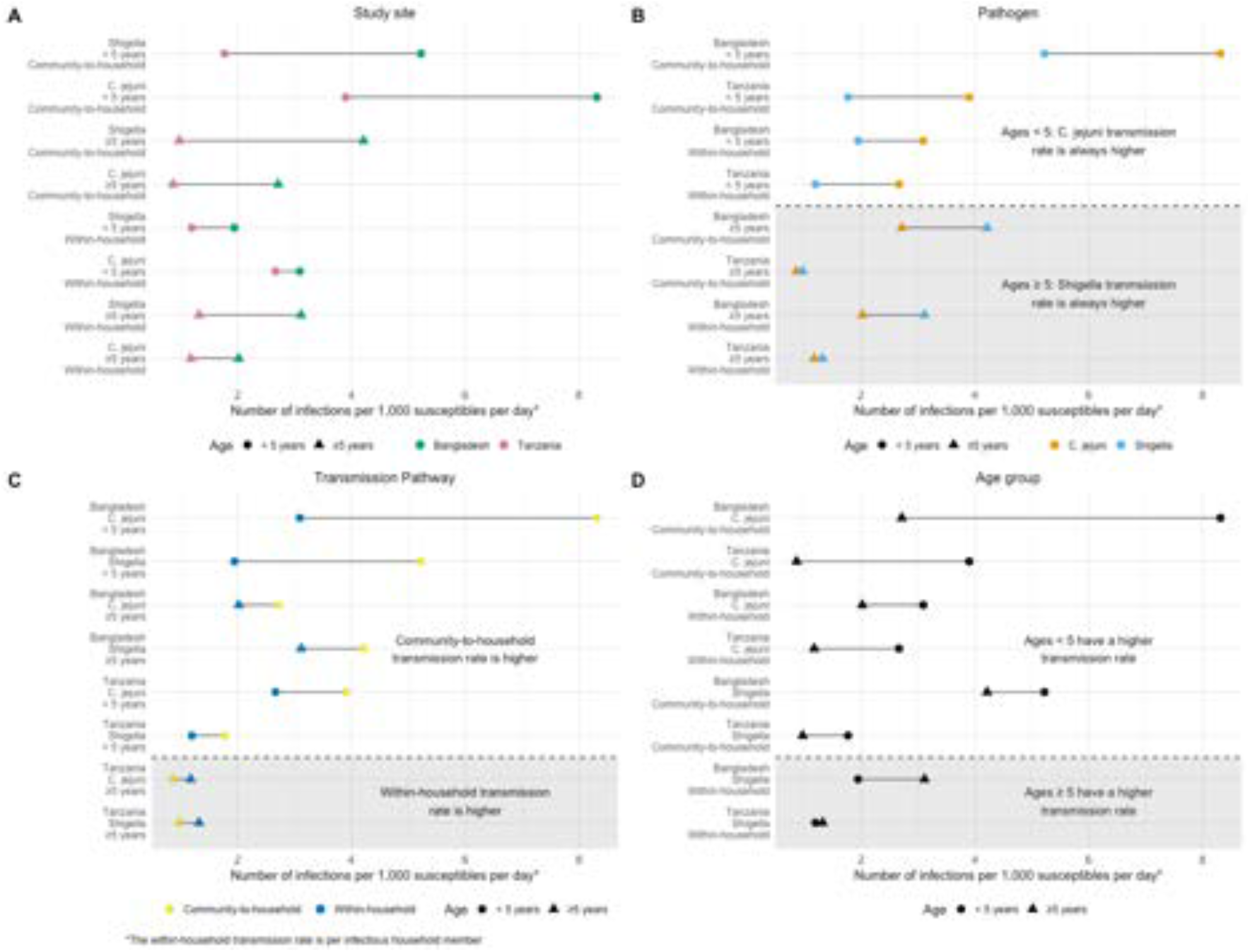
Paired lollipop plots showing the median posterior site-, pathogen, pathway- and age-specific transmission rates from the full, forward-selected model compared across covariate pairs matched by (A) study site, (B) pathogen, (C) transmission pathway, and (D) age group. Each panel shows the same 16 transmission rate parameters rearranged according to the covariate of interest. Panel (A) shows that Bangladesh (green) always has a higher transmission rate than Tanzania (pink). Panel (B) shows that *C. jejuni* transmission rates (orange) were always higher for children <5 while *Shigella* transmission rates (light blue) were always higher for those ≥5 years. Panel (C) shows that the community-to-household transmission rate (yellow) was higher than the household transmission rate (dark blue) except in the older groups in Tanzania where transmission is the lowest. Finally, panel (D) shows that children <5 (circles) always had a higher transmission rate than those ≥5 years (triangles) except for within-household *Shigella* transmission.

The pathogen–pathway interaction term was not selected into the model, as adding it did not improve the median model BIC (indeed, it did not improve the BIC in *any* of the simulated datasets; median difference in BIC: −8.59; range: −19.18 to −0.38). This result indicates that, after accounting for age and study site, and relevant two-way interactions, the relative contribution of community-to-household and within-household transmission did not differ between *Shigella* and *Campylobacter*. Given that this pathogen– pathway interaction was our primary research question, we estimated the magnitude of the interaction term in the maximal model also including this term. The rate ratio estimate was 0.98 (95% CrI: 0.80– 1.54), and the other rate ratios were similar to the final forward-selected model (see Supplemental Table S2), generating similar transmission rate estimates to the main analysis (see Supplemental Figure S5).

The results from the sensitivity analyses using alternative Ct cutoffs of 35 and 27 were similar to the main analysis (see Supplemental Figure S6 and S7). All the results were qualitatively the same as the main analysis (e.g., the transmission rate was always higher in Bangladesh compared to the analogous transmission rate in Tanzania), although specific estimates differed quantitatively (particularly because a different Ct cutoff translates to a different number of infections overall and thus necessarily changed the transmission rates).

To relax the assumption that an individual was infectious continuously between two successive positive PCR tests, we conducted sensitivity analyses using alternative infection durations. Most qualitative comparisons from Figure 3 were preserved (see Supplemental Figures S8–11). However, some comparisons were sensitive to relaxing this assumption. While the primary analysis estimated higher within-household transmission for *C. jejuni* among children <5 in Bangladesh than Tanzania, all alternative infection-duration models reversed this ordering, estimating instead higher transmission rates in Tanzania. The pathogen-specific ordering among individuals ≥5 years in Tanzania was also sensitive to infection duration assumptions: although the primary analysis estimated higher *Shigella* than *C. jejuni* transmission for both community-to-household and within-household transmission, this ordering was reversed under several alternative infection-duration models (Figure S9). The age-specific ordering of within-household *Shigella* transmission in Tanzania was similarly reversed under some alternative assumptions (Figure S11). Thus, while some specific comparisons depended on assumptions about infection duration, the broader findings of substantial effect modification by age and the greater relative contribution of community-to-household transmission among children <5 were preserved.

## Discussion

In this work, we leveraged longitudinally collected household stool samples to estimate the importance of within-household and community-to-household transmission for *Shigella* and *Campylobacter*. Relative to Tanzania, the incidence rate of *Campylobacter* was 1.4 times higher in Bangladesh, and the incidence rate of *Shigella* was 2.3 times higher. We estimated low levels of symptomatic infections (1–25%), consistent with the broader literature showing that asymptomatic infections are common.^18,19^ Direct comparison of these estimates with previous studies is challenging because the vast majority of papers that examined asymptomatic carriage report the proportion of asymptomatic stool samples that test positive for a pathogen, as opposed to the proportion of infections that were asymptomatic, which we report here. In our study, 16.9% and 8.7% of asymptomatic stools in children <5 were positive for *Shigella* in Bangladesh and Tanzania, respectively, compared with 13% and 18% of non-diarrheal stools in previous longitudinal surveillance from the same settings.^20^ For *Campylobacter*, 34.8% and 20.3% of asymptomatic stools were positive in Bangladesh and Tanzania, respectively, similarly demonstrating the high prevalence of asymptomatic detection reported in previous longitudinal studies in these settings.^21^ We estimated median durations of PCR positivity of 29 days for *Shigella* and 32 days for *Campylobacter*. Although estimates of pathogen persistence vary across studies because of differences in study design and population, our findings are broadly consistent with previous longitudinal studies reporting duration of PCR positivity for *Campylobacter*, which reported mean infection durations ranging from 30–53 days. ^22–24^

In the vast literature on the transmission routes of enteric pathogens, differences between the epidemiology of *Shigella* and *Campylobacter* have typically been explained by pathogen biology. Relative to *Shigella, Campylobacter* has high environmental persistence, making it more prone to environmentally mediated infection pathways such as contaminated food or water.^8–13^ In contrast, *Shigella* has a lower environmental persistence and the literature typically attributes outbreaks to person-to-person and household transmission.^4–7^ Consistent with this literature, in our preliminary analysis, we found that the ratio of community-to-household to the within-household transmission rates was more than twofold higher for *Campylobacter* than *Shigella*. However, our forward-selected final model suggested that the apparent difference between pathogens was largely explained by effect modification by age, rather than an interaction between pathogen and pathway.

Specifically, we found that relative to individuals aged ≥5 years, children <5 had greater *Campylobacter* transmission rates compared to *Shigella*, and higher relative community-to-household transmission (compared to within-household) rates, regardless of pathogen. Even within the household, children <5 had higher transmission rates than household members ≥5 years for *Campylobacter* (Figure 3D). In contrast, in the main analysis, older children and adults had higher transmission rates for *Shigella* than *Campylobacter* (Figure 3B). This differential was most apparent in Bangladesh (Figure 3C). As a result, the greater contribution of community-to-household transmission to *Campylobacter* identified in the preliminary analysis was largely attributable to the age distribution of infections and differences in transmission pathway by age. These findings indicate that the dominant transmission pathways observed at the population level for each pathogen can depend substantially on age, likely driven by a combination of differences in behavior, exposure, and susceptibility. Consistent with the importance of infection sources not attributable to infectious household members among young children, Singh et al. identified chickens as the primary reservoir of *C. jejuni* infection in infants in rural Ethiopia, also finding that mothers and siblings could act as intermediate sources.^25^ Although *Shigella* and *Campylobacter* have biological differences that may favor different transmission routes, these findings demonstrate that population-level transmission patterns can vary substantially across age groups and cannot be inferred from pathogen biology alone.

Our work has several strengths. The monthly stool samples collected among a cohort of household members allowed us to longitudinally assess household transmission, accounting for asymptomatic shedding and infection. Leveraging these data in household transmission model allowed us to account for the role of asymptomatic infections in household transmission chains and subsequently directly quantify the relative contribution of infection attributable to infectious household members versus other sources for two important pathogens. Our work also has limitations. Quantitative PCR is highly sensitive to the presence of bacterial DNA which may influence infection classification. To address this possibility, we conducted a sensitivity analysis using a more stringent Ct cutoff (27) and a less stringent cutoff (35) to assess whether our conclusions changed. Given that the results were stable between these analyses, we expect this limitation to only minimally impact the results. Our model also does not account for the possibility of multiple household infections occurring from the same community exposure in close succession. The imputed infection timing could make these infections appear sequential, potentially attributing external acquisition to within-household transmission and inflating the estimated within-household transmission rate. Finally, although transmission patterns may vary substantially across early childhood, particularly in children <5, the relatively small number of children aged 1–5 years in the cohorts (n = 119 in Tanzania and n = 35 in Bangladesh) precluded a more finely age-stratified analysis within children <5.

Pathogens have important biological differences that determine how successfully they are transmitted along various pathways; however, our results, which demonstrated that transmission patterns varied substantially by age group, reinforce the idea that infection is a product of pathogen, host, and exposure factors. After accounting for age-specific heterogeneity in transmission rates, we found no evidence that the relative contribution of within-household versus community-to-household transmission differed between *Shigella* and *Campylobacter*. The specific transmission pathways underlying these components—including person-to-person, environmental, food- or waterborne, and zoonotic transmission—may nevertheless differ between pathogens and populations. Analysis of the contemporaneous environmental and animal sampling data collected in this study will later allow us to determine whether these pathways contribute differentially to external acquisition for each pathogen, particularly among young children for whom this transmission component dominates.

## Data Availability

All data produced in the present study are available upon reasonable request to the authors.

## References

1. Diarrhoeal disease. Accessed August 18, 2025. https://www.who.int/news-room/fact-sheets/detail/diarrhoeal-disease

2. Grembi JA, Lin A, Karim MA, et al. Effect of Water, Sanitation, Handwashing, and Nutrition Interventions on Enteropathogens in Children 14 Months Old: A Cluster-Randomized Controlled Trial in Rural Bangladesh. J Infect Dis. 2023;227(3):434–447. doi:10.1093/infdis/jiaa549

3. Lin A, Ercumen A, Benjamin-Chung J, et al. Effects of Water, Sanitation, Handwashing, and Nutritional Interventions on Child Enteric Protozoan Infections in Rural Bangladesh: A Cluster-Randomized Controlled Trial. Clin Infect Dis. 2018;67(10):1515–1522. doi:10.1093/cid/ciy320

4. Hald T, Aspinall W, Devleesschauwer B, et al. World Health Organization Estimates of the Relative Contributions of Food to the Burden of Disease Due to Selected Foodborne Hazards: A Structured Expert Elicitation. PloS One. 2016;11(1):e0145839. doi:10.1371/journal.pone.0145839

5. Butler AJ, Thomas MK, Pintar KDM. Expert elicitation as a means to attribute 28 enteric pathogens to foodborne, waterborne, animal contact, and person-to-person transmission routes in Canada. Foodborne Pathog Dis. 2015;12(4):335–344. doi:10.1089/fpd.2014.1856

6. Vally H, Glass K, Ford L, et al. Proportion of illness acquired by foodborne transmission for nine enteric pathogens in Australia: an expert elicitation. Foodborne Pathog Dis. 2014;11(9):727–733. doi:10.1089/fpd.2014.1746

7. George CM, Ahmed S, Talukder KA, et al. Shigella Infections in Household Contacts of Pediatric Shigellosis Patients in Rural Bangladesh. Emerg Infect Dis. 2015;21(11):2006–2013. doi:10.3201/eid2111.150333

8. McLure A, Smith JJ, Firestone SM, et al. Source attribution of campylobacteriosis in Australia, 2017-2019. Risk Anal Off Publ Soc Risk Anal. 2023;43(12):2527–2548. doi:10.1111/risa.14138

9. Lake RJ, Campbell DM, Hathaway SC, et al. Source attributed case-control study of campylobacteriosis in New Zealand. Int J Infect Dis IJID Off Publ Int Soc Infect Dis. 2021;103:268–277. doi:10.1016/j.ijid.2020.11.167

10. Rosner BM, Schielke A, Didelot X, et al. A combined case-control and molecular source attribution study of human Campylobacter infections in Germany, 2011–2014. Sci Rep. 2017;7(1):5139. doi:10.1038/s41598-017-05227-x

11. Wagenaar JA, French NP, Havelaar AH. Preventing Campylobacter at the Source: Why Is It So Difficult? Clin Infect Dis. 2013;57(11):1600–1606. doi:10.1093/cid/cit555

12. Boysen L, Rosenquist H, Larsson JT, et al. Source attribution of human campylobacteriosis in Denmark. Epidemiol Infect. 2014;142(8):1599–1608. doi:10.1017/S0950268813002719

13. Cody AJ, Maiden MC, Strachan NJ, McCarthy ND. A systematic review of source attribution of human campylobacteriosis using multilocus sequence typing. Euro Surveill Bull Eur Sur Mal Transm Eur Commun Dis Bull. 2019;24(43):1800696. doi:10.2807/1560-7917.ES.2019.24.43.1800696

14. Mbelele PM, Katengu S, Wettstone EG, et al. Sources and transmission dynamics of Campylobacter and Shigella using culture-independent molecular methods in Tanzania (SCAT): a protocol for a prospective observational cohort study. medRxiv. Preprint posted online June 25, 2026:2026.06.23.26356359. doi:10.64898/2026.06.23.26356359

15. Fischer GH, Hashmi MF, Paterek E. Campylobacter Infection. In: StatPearls. StatPearls Publishing; 2025. Accessed March 24, 2025. http://www.ncbi.nlm.nih.gov/books/NBK537033/

16. Awofisayo-Okuyelu A, Hall I, Adak G, HAwker JI, ABbott S, McCARTHY N. A systematic review and meta-analysis on the incubation period of Campylobacteriosis. Epidemiol Infect. 2017;145(11):2241–2253. doi:10.1017/S0950268817001303

17. Chai SJ, Gu W, O’Connor KA, Richardson LC, Tauxe RV. Incubation periods of enteric illnesses in foodborne outbreaks, United States, 1998–2013. Epidemiol Infect. 2019;147:e285. doi:10.1017/S0950268819001651

18. Del Carpio-Orantes L, Sánchez-Díaz JS, Peniche Moguel KG, et al. 718. Asymptomatic campylobacteriosis as a risk factor for developing acute neurological syndromes in Veracruz, Mexico. Open Forum Infect Dis. 2020;7(Supplement_1):S410. doi:10.1093/ofid/ofaa439.910

19. Guerrero L, Calva JJ, Morrow AL, et al. Asymptomatic Shigella infections in a cohort of Mexican children younger than two years of age. Pediatr Infect Dis J. 1994;13(7):597–602. doi:10.1097/00006454-199407000-00003

20. Rogawski McQuade ET, Shaheen F, Kabir F, et al. Epidemiology of Shigella infections and diarrhea in the first two years of life using culture-independent diagnostics in 8 low-resource settings. PLoS Negl Trop Dis. 2020;14(8):e0008536. doi:10.1371/journal.pntd.0008536

21. Amour C, Gratz J, Mduma E, et al. Epidemiology and Impact of Campylobacter Infection in Children in 8 Low-Resource Settings: Results From the MAL-ED Study. Clin Infect Dis Off Publ Infect Dis Soc Am. 2016;63(9):1171–1179. doi:10.1093/cid/ciw542

22. Amin AB, Garcia Quesada M, Liu J, et al. Natural history parameters for enteric pathogens to inform modeling studies of diarrhea among children in low-resource settings: results from the MAL-ED longitudinal birth cohort. BMC Infect Dis. 2025;26(1):106. doi:10.1186/s12879-025-12265-8

23. Chen D, Havelaar AH, Platts-Mills JA, Yang Y. Acquisition and clearance dynamics of Campylobacter spp. in children in low- and middle-income countries. Epidemics. 2024;46:100749. doi:10.1016/j.epidem.2024.100749

24. McCartney NK, Thilakarathna SH, Hluchy T, Pillai DR, Chui L, Berenger BM. Bacterial gastroenteritis in the world of culture-independent diagnostic testing: a study to evaluate the kinetics of bacterial shedding by culture and CIDT. Microbiol Spectr. 13(7):e00227–25. doi:10.1128/spectrum.00227-25

25. Singh N, Thystrup CAN, Hassen BM, et al. Transmission pathways of Campylobacter jejuni between humans and livestock in rural Ethiopia are highly complex and interdependent. Gut Pathog. 2025;17:26. doi:10.1186/s13099-025-00691-7

