## Supplemental Material for "Comparative analysis of *Shigella* and *Campylobacter* transmission in paired longitudinal household cohort studies in urban Bangladesh and rural Tanzania"

### Supplementary appendix

Table of Contents

*1. Data augmentation* ... *2*

1.1. Data augmentation: Step 1 ... 5

1.2. Length of infection ... 7

1.3. Infection start date ... 8

*2. Household model likelihood* ... *9*

*3. Age-unadjusted model results* ... *10*

*4. Model rate ratios* ... *11*

*5. Transmission rate parameters from forward-selected model* ... *12*

*6. Sensitivity analysis with six interaction terms* ... *13*

*7. Sensitivity analysis with a Ct cutoff of 35* ... *14*

*8. Sensitivity analysis with a Ct cutoff of 27* ... *15*

*9. Sensitivity analysis of infection duration assumptions* ... *16*

#### 1. Data augmentation

We defined an infectiousness vector for each study participant $i$ spanning each day $d \in \{1, \dots, m_i\}$ of their enrollment period. This vector, $v_i = (v_{i,1}, v_{i,2}, \dots, v_{i,m_i})$, could take on one of 3 values: $-$ if individual $i$ had Ct>30 on day $d$, + if individual $i$ had Ct≤30 on day $d$, and 0 (unknown status) otherwise.

Given stool samples were only collected twice weekly to monthly, the infectious status is unknown (0) on most days in the original data. We multiply imputed the days with missing infectious statuses to create 10,000 augmented datasets for subsequent analysis. Augmentation was done in four steps (see Figure S1 for an illustrated example):

Step 1: If two consecutive tests indicated positive infectious status, we assumed that the individual was also infectious each day between that pair of tests. We treated each resulting set of consecutive infectious days and adjacent sequences of days with an unknown infection status as a single infection with an unknown start and end date (see *Section 1.1*).

Step 2: To estimate the start and end dates for each discrete infection, we assigned each infection an infection length $l$ greater than or equal to the current length (see *Section 1.2*) and an infection start date $s$ (see *Section 1.3*).

Step 3: We prepended a latent period of 3 days to each augmented infection, corresponding to the period where the individual had been infected with the pathogen but was not yet infectious.

Step 4: An individual was assumed to be uninfected on days not determined to be infectious or latent in steps 1–3.

As the data augmentation algorithm is stochastic (i.e., the estimated infection length $l$ and start date $s$ are random variables), we generated 10,000 data augmentation iterates and applied the household model to each iterate, averaging over the multiple realizations. We denote the $k^{\text{th}}$ iteration of the augmented daily infectious status for individual $i_j$ in household $j$ by $\hat{v}_{j,i_j,k}$.

Household-level augmented data was generated by concatenating the infection vectors across all individuals in household $j$ to create a household infection matrix, $A^{j,k} = [\hat{v}_{j,1,k}, \hat{v}_{j,2,k}, \dots, \hat{v}_{j,n_j,k}]$ where $n_j$ is the number of people in household $j$. A plot showing the density of 1,000 data augmentations in the four example households can be found in Figure S2.

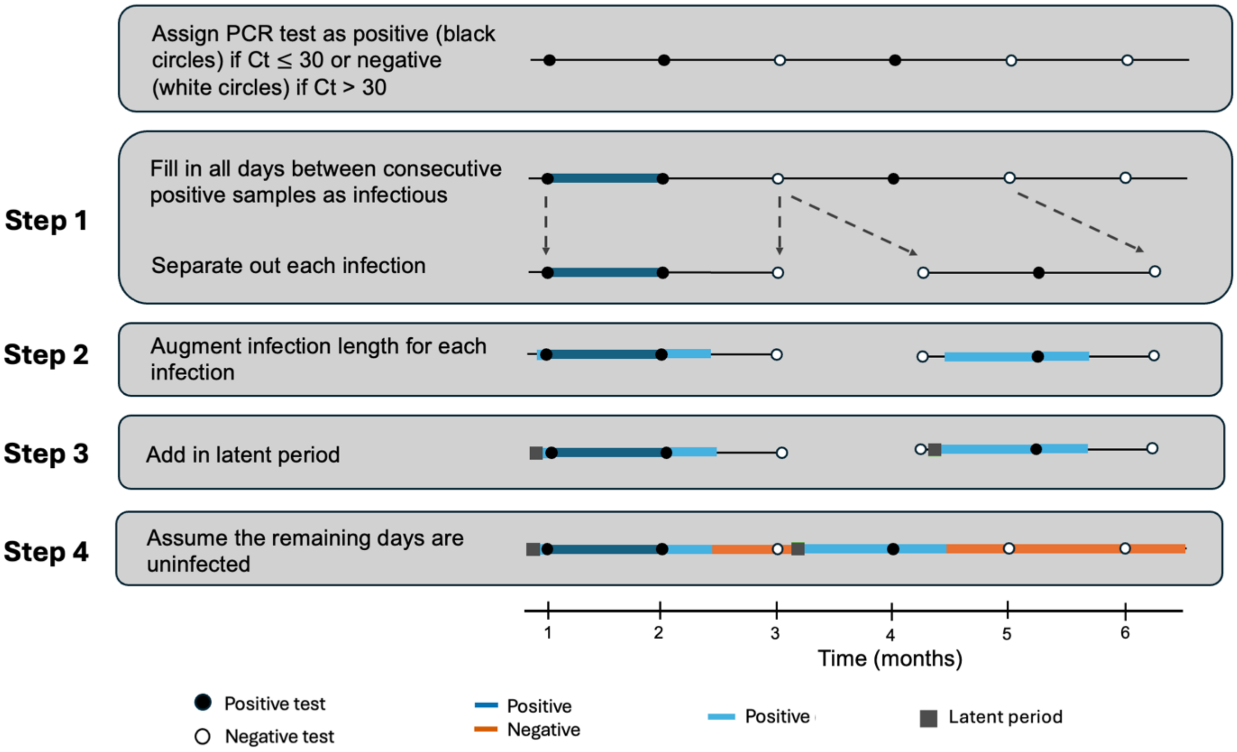


**Figure S1:** Example of the data augmentation process for one individual with six months of data. Each individual was determined to be infectious or not for a given pathogen on study visit days if their stool sample had a PCR Ct of ≤ 30 or >30, respectively. In the first step, dates between consecutive positive tests were assumed to be positive (dark blue lines). Individual infections were separated for data augmentation according to their maximum possible length (see Supplemental Material, *Section 1.1*). In the second step, infection lengths were increased by a value pulled from an empirical infection length distribution (see Figure S3), and the infection was set to begin at a starting date randomly selected from a uniform distribution (see Supplemental Material, *Section 1.3*). In step three, each infection was prepended by a three-day latent period. Finally, in step four, all days that were not determined to be infectious or latent were assumed to be uninfected.

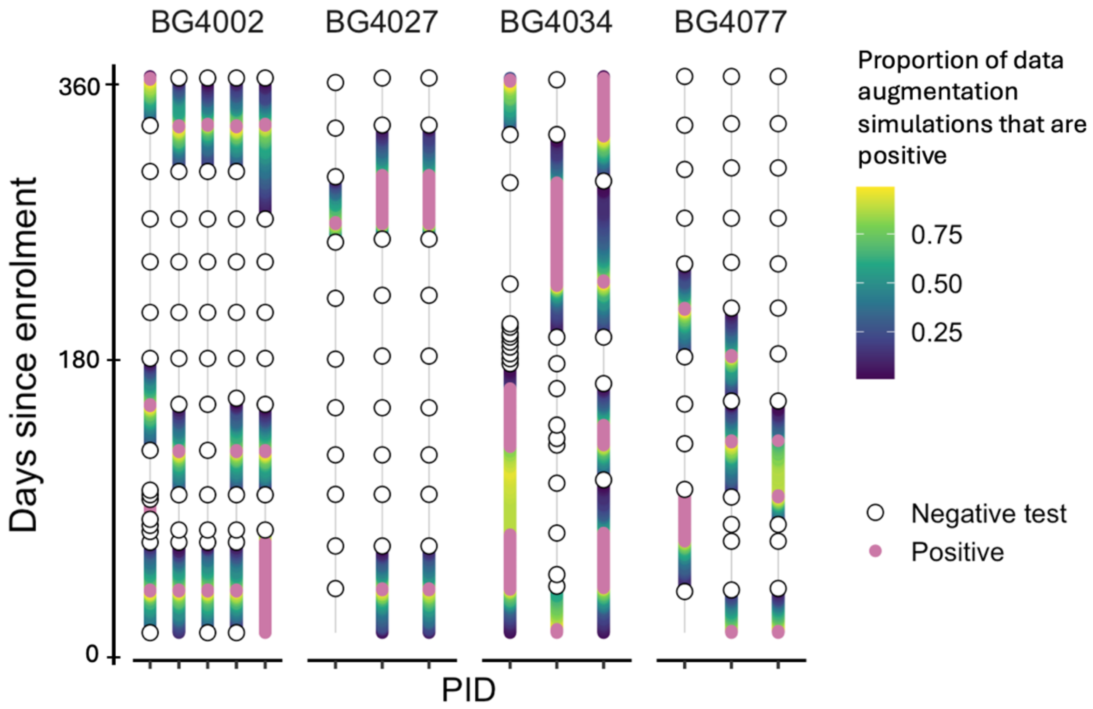


**Figure S2:** Density plots of augmented data across 1,000 simulations. Pink corresponds to dates that were positive from Step 1, and blue-yellow colored data are dates that were augmented to be positive across 1,000 simulations, with yellow corresponding to a higher proportion of simulations being positive on that day and blue low.

##### 1.1. Data augmentation: Step 1

For an individual $i$, the vector $v_i = (v_{i,1}, v_{i,2}, \ldots, v_{i,m_i})$ is defined by:

$$v_{i,k} = \begin{cases} - \text{ if individual } i \text{ tested negative on day } k \text{ (Ct} > 30) \\ + \text{ if individual } i \text{ tested positive on day } k \text{ (Ct } \leq 30) \\ \quad 0 \text{ otherwise (unknown infection status)} \end{cases}$$

And $m_i$ is the number of days the participant was enrolled in the study. We fill in all the days between consecutive positive tests as infectious. If the two consecutive positive tests are greater than 30 days apart (i.e., a monthly test was missing), we leave the infection status as unknown.

```
ALGORITHM 1: INFERRING DATES OF POSITIVITY

set vec = v_i , the daily infection status for individual i
set last_positive_test = NULL
for k in length(v_i):
    if vec[k] == +:
        if k – last_positive_test =< 30 and last_positive_test != NULL:
            set all dates between last_positive_test and k in vec to +
        update last_positive_test = k
return vec
```

To get the distribution for the length of infection and perform data augmentation, we need to parse out infections for each individual such that the dates of infection (i.e., $v_{i,k} = +$), and all immediately surrounding dates that are of unknown infection status (i.e., $v_{i,k} = 0$) are included. This is equivalent to pulling out the dates between the nearest two negative tests. This process is outlined in Algorithm 2.

```
ALGORITHM 2: SEPARATING OUT INFECTIONS

set vec = v_i , the daily infection status for individual i
set result = [] # indices of infection
while i <= length(vec):

    # PART A: identifying indices of infectious days

    if vec[i] = +:
        set start = i
        while (i < length(vec) & vec[i] = +): # while individual still infectious
            i = i + 1
        set end = i – 1

        # PART B: identifying consecutive days that are unknown infection status
        # set surrounding dates to start and end, then update if surrounding dates
        # are unknown infection status (0)

        set start_surround = start
        set end_surround = end
        if start > 1 and vec[start – 1] = 0:
```

```
            while(start_surround > 1 and vector[start_surround – 1] == 0:
                start_surround = start_surround – 1
        if end < n and vec[end_surround + 1] = 0:
            while end_surround < n and vector[end_surround + 1] == 0:
                end_surround = end_surround + 1
        append start surround and end_surround to result
    i = i + 1
return result
```

The resulting vector is the indices of dates that each person is infectious and the surrounding unknown dates.

##### 1.2. Length of infection

To augment the infection data from Step 1, we drew numbers corresponding to the length of infection and the start date of the infection. We randomly pulled the infection length $l$ from an empirically estimated infection length density distribution using the infections from Step 1. Because the true infection length must be greater than or equal to the current infection length $(a)$ and less than the number of days between the nearest two negative tests $(b)$, the augmented length for an infection $l$ was drawn conditionally so that $a \leq l \leq b$. We drew the date the infection started $s$ from a uniform distribution $s \sim \text{Uniform}\{p_{\text{start}}, p_{\text{end}}\}$, where the earliest and latest possible start dates, $p_{\text{start}}$ and $p_{\text{end}}$ were given by Eq. S1 and S2 in the Supplemental Material (see *Section 1.3*).

We empirically estimate a distribution of the length of infection for each pathogen. For each infection across all the individuals in the study, we draw 1,000 potential infection durations from a uniform distribution, with the minimum length set to the infection duration from Step 1 and the maximum possible length equal to the number of days between the most recent negative test and the following negative test. From the drawn infection lengths, we generate an empirical infection length distribution for each pathogen (see Figure S3). The algorithm for this process is described below.

ALGORITHM 3: DISTRIBUTION OF INFECTION LENGTH

set empty vector $L = []$ # vector of infection lengths
for individual $k$ in K:
    set E to be the parsed out infections according to Algorithm 2
    for $e_j$ in $E$:
        set length of vector $n = \text{length}(e_j)$ # maximum possible length of infection
        set sum of vector $l = \sum e_j$ # minimum possible length of infection
        generate 1,000 random draws from a uniform distribution $x \sim \text{Uniform}\{l, n\}$
        concatenate $x$ to $L$

generate 2-D density function of $L$

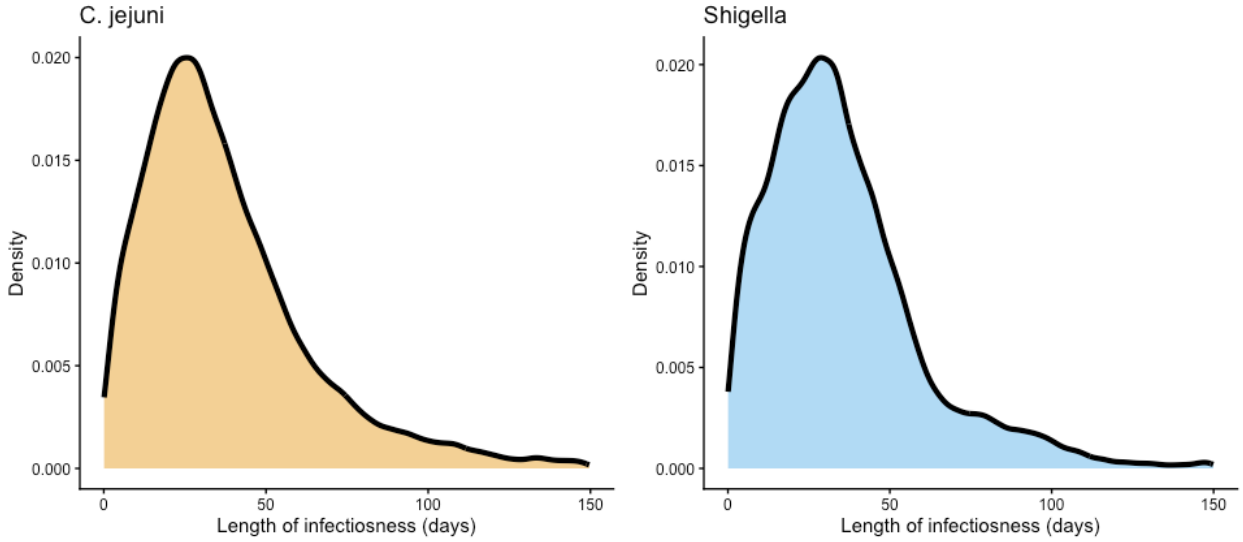


**Figure S3:** Density distribution of infection length (days) for *C. jejuni* and *Shigella*.

##### 1.3. Infection start date

Given a single infection $v_j$ with the following properties,

| Variable | Definition | Value |
|---|---|---|
| $\boldsymbol{n}$ | Length of $v_j$ | $\lvert v_j \rvert$ |
| $\boldsymbol{l_c}$ | Length of the infection | $\sum v_j$ |
| $\boldsymbol{i}_{\text{start}}$ | Index of the first day of infection | $\min(i \in \{1,2,\ldots n\} \mid v_j = 1)$ |
| $\boldsymbol{i}_{\text{end}}$ | Index of the last day of infection | $\max(i \in \{1,2,\ldots n\} \mid v_j = 1)$ |

The augmented length of infection $l$ is pulled from the respective distribution in Figure S3 where $a = l_c$ and $b = n$. The infection can be positioned anywhere in $v_j$ such that $\sum \hat{v}_j = l$ (i.e., all of the days previously determined to be infectious in $v_j$ remain infectious). The earliest and latest allowable infection start indices are therefore

$$\mathrm{s}_{\min} = \min(i_{\text{start}}, n - l) \qquad (\text{Eq.}\,S1)$$

and

$$\mathrm{s}_{\max} = \max(i_{\text{end}} - l, 0) \qquad (\text{Eq.}\,S2)$$

The first date of infection is pulled from $s \sim \text{Uniform}\{\mathrm{s}_{\min}, \mathrm{s}_{\max}\}$. The final infection vector is defined as:

$$\hat{v}_{j,k} = \begin{cases} + \text{ for } k \in [s, s + l) \\ - \text{ otherwise} \end{cases}$$

**2. Household model likelihood**

In each augmented dataset for each individual $i_j$ in household $j$, $A^{j,k}$, we defined vector $t^{*}_{j,i_j,k}$ as their imputed day of infection (the first day of the latent period for each infection) and $t^{**}_{j,i_j,k}$ as the imputed days they were not infectious. The individual-level log-likelihood has the following expression:

$$\log(\Pr\left(t^{*}_{j,i_j,k}\right)) = \sum_{t\in t^{*}_{j,i_j,k}} \log\left(1-\exp(-\lambda(t)_{j,i_j,k})\right) + \sum_{t\in t^{**}_{j,i_j,k}} \log\left(\exp(-\lambda(t)_{j,i_j,k})\right), \quad (\text{Eq.} S3)$$

Where $\lambda(t)_{j,i_j,k}$ is the daily force of infection experienced by individual $i_j$ in household $j$, defined in Eq. 2 in the main text). This term represents the sum of the logs of the probability of individual $i_j$ getting infected on the day they got infected and of the probability of avoiding infection on the days they were not infected. The overall log-likelihood for data augmentation iterate $k$ is then given by the sum of all individual likelihoods in household $j$ summed over all households ($J$):

$$\mathcal{L}_k = \sum_{j\in J}\sum_{i_j\in j} \log\left(Pr\left(t^{*}_{j,i_j,k}\right)\right) \quad (\text{Eq.} S4)$$

Through the parameterization in Eqs 1 and 2 in the main text, the covariate log rate ratios are connected to the model likelihood.

#### 3. Age-unadjusted model results

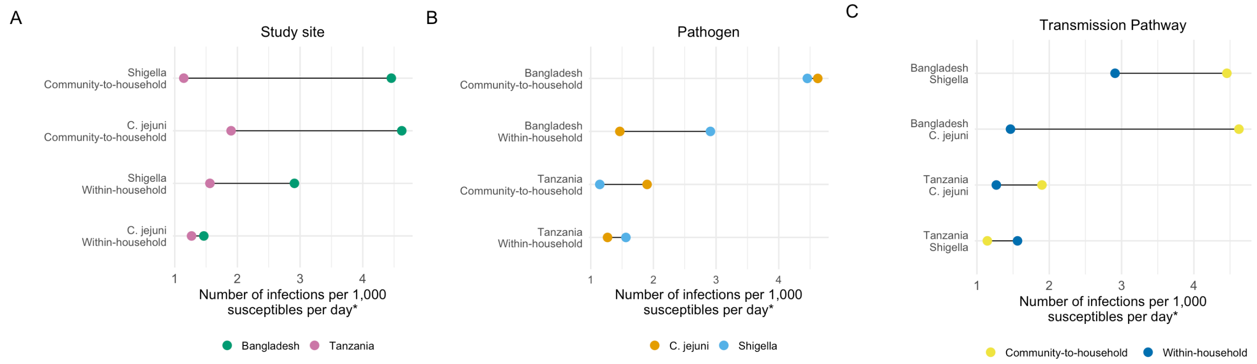


**Figure S4:** Paired lollipop plots showing the median posterior site-, pathogen, and pathway-specific transmission rates compared across covariate pairs matched by (A) study site, (B) pathogen, and (C) transmission pathway in the age-unadjusted model. Each panel shows the same eight transmission rate parameters rearranged according to the covariate of interest. Panel (A) shows that Bangladesh (green) always had higher transmission rates than Tanzania (pink), (B) shows that *C. jejuni* (orange) had a higher community-to-household transmission rate than *Shigella* (light blue), while *Shigella* had a higher within-household transmission rate than *C. jejuni*. Panel (C) shows that the community-to-household transmission rate (yellow) was higher than the within-household transmission rate (blue) for *C. jejuni,* and lower for *Shigella*.

**Supplemental Table S1:** Transmission rate parameters in age-unadjusted model

| Country | Pathogen | Community-to-household transmission rate* | Within-household transmission rate* | Ratio of community-to-household to within-household transmission rates |
|---|---|---|---|---|
| Bangladesh | *Shigella* | 4.46 (4.33, 4.58) | 2.91 (2.63, 3.22) | 1.53 (1.35, 1.74) |
| | *Campylobacter* | 4.62 (4.50, 4.75) | 1.46 (1.25, 1.70) | 3.15 (2.66, 3.78) |
| Tanzania | *Shigella* | 1.14 (1.07, 1.19) | 1.56 (1.40, 1.80) | 0.73 (0.60, 0.84) |
| | *Campylobacter* | 1.90 (1.81, 1.99) | 1.27 (1.11, 1.42) | 1.51 (1.29, 1.77) |

*Number of infections per 1,000 susceptibles per day. Within-household transmission rates are per infectious household member

**4. Model rate ratios**

**Supplemental Table S2:** Rate ratio estimates for the main analysis with 5 interaction terms, the model including the sixth pathway by pathogen interaction term, and the preliminary model.

| | **Forward-selected model (Ct=30)** | **Model including pathway x pathogen** | **Preliminary model** |
|---|---|---|---|
| Main effects | | | |
| Intercept | 1.32 e -3 (1.12, 1.49) | 1.34 e -3 (1.12, 1.66) | 1.56 e -3 (1.40, 1.80) |
| Study site [ref: Tanzania] | 2.34 (2.01, 2.64) | 2.37 (1.97, 2.77) | 1.87 (1.61, 2.13) |
| Pathogen [ref: Shigella] | 0.89 (0.84, 0.95) | 0.89 (0.65, 1.04) | 0.80 (0.71, 0.94) |
| Age [ref: 5+] | 0.92 (0.76, 1.09) | 0.86 (0.64, 1.16) | — |
| Pathway [ref: Household] | 0.73 (0.61, 0.86) | 0.74 (0.55, 0.89) | 0.73 (0.60, 0.84) |
| Interaction terms | | | |
| Study site x pathogen | 0.73 (0.66, 0.77) | 0.72 (0.62, 0.80) | 0.62 (0.60, 0.64) |
| Study site x age | 0.68 (0.64, 0.73) | 0.69 (0.64, 0.73) | — |
| Study site x pathway | 1.86 (1.54, 2.24) | 1.83 (1.53, 2.28) | 2.09 (1.75, 2.55) |
| Pathogen x age | 2.46 (2.38, 2.65) | 2.51 (2.29, 2.69) | — |
| Pathway x age | 1.97 (1.60, 2.53) | 2.11 (1.52, 2.85) | — |
| Pathway x pathogen | — | 0.98 (0.80, 1.54) | 2.05 (1.69, 2.49) |
| BIC | 27268 (27146, 27474) | 27279 (27156, 27483) | 27470 (27437, 27755) |

##### 5. Transmission rate parameters from forward-selected model

**Supplemental Table S3:** Transmission rate estimates for the 16 possible combinations of study site, pathogen, age, and transmission pathway.

| Study site | Pathogen | Age | Transmission pathway | Transmission rate (infections per 1,000 susceptible per day) |
|---|---|---|---|---|
| Bangladesh | *Shigella* | < 5 | Community-to-household | 5.22 (5.08, 5.35) |
| | | | Within-household* | 1.94 (1.62, 2.23) |
| | | ≥ 5 | Community-to-household | 4.21 (4.08, 4.36) |
| | | | Within-household* | 3.11 (2.77, 3.42) |
| | *Campylobacter* | < 5 | Community-to-household | 8.31 (8.07, 8.52) |
| | | | Within-household* | 3.09 (2.57, 3.55) |
| | | ≥ 5 | Community-to-household | 2.71 (2.60, 2.83) |
| | | | Within-household* | 2.02 (1.78, 2.17) |
| Tanzania | *Shigella* | < 5 | Community-to-household | 1.77 (1.67, 1.83) |
| | | | Within-household* | 1.19 (1.03, 1.47) |
| | | ≥ 5 | Community-to-household | 0.97 (0.90, 1.03) |
| | | | Within-household* | 1.32 (1.19, 1.48) |
| | *Campylobacter* | < 5 | Community-to-household | 3.90 (3.59, 4.10) |
| | | | Within-household* | 2.66 (2.32, 3.19) |
| | | ≥ 5 | Community-to-household | 0.86 (0.79, 0.95) |
| | | | Within-household* | 1.17 (1.08, 1.31) |

*Within-household transmission rates are per infectious household member

#### 6. Sensitivity analysis with six interaction terms

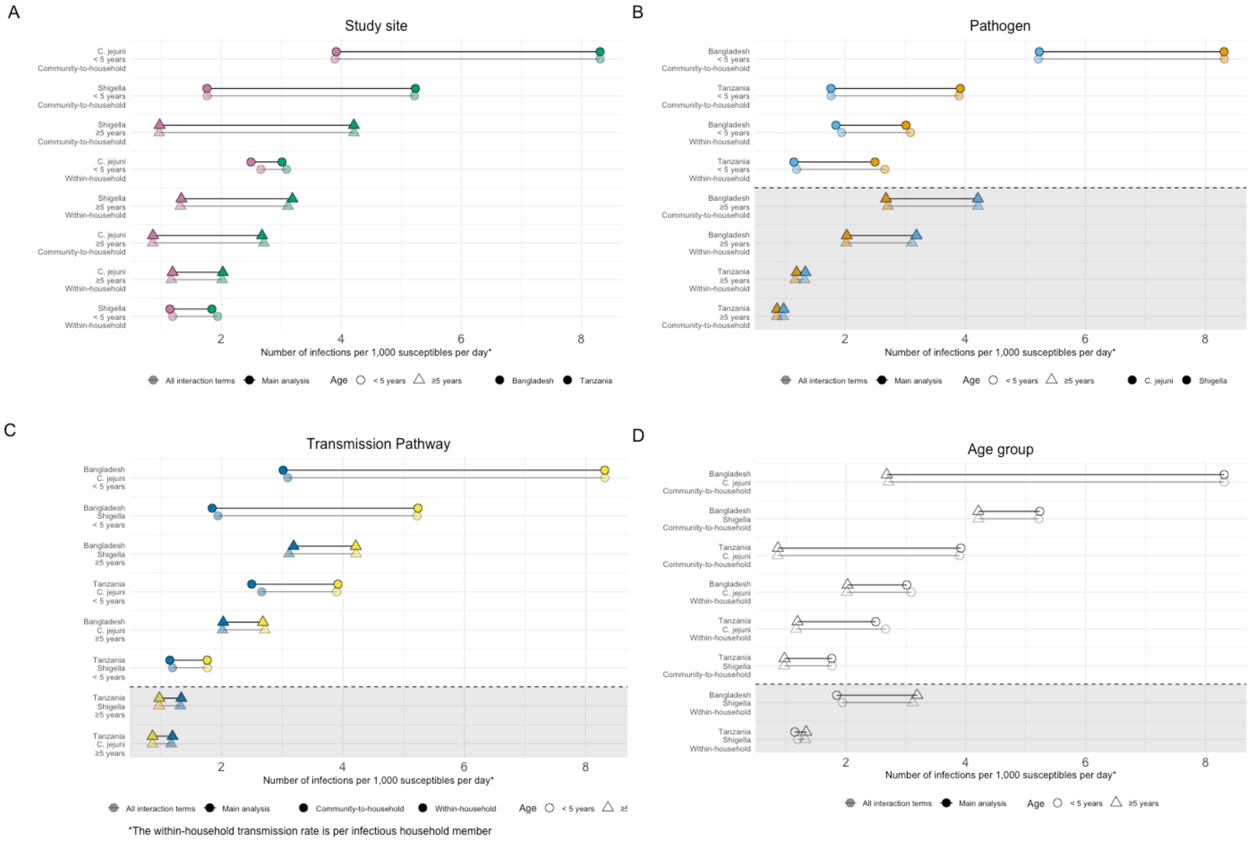


**Figure S5:** Sensitivity analysis showing the results from the main analysis (opaque) compared to the sensitivity analysis with all six interaction terms (translucent). Lollipop plots showing the median posterior value for each transmission rate parameter compared across pairs matched by study site (A), pathogen (B), transmission pathway (C), and age group (D). Each panel shows the same 16 transmission rate parameters rearranged according to the covariate of interest. Panel (A) shows that Bangladesh (green) always has a higher transmission rate than Tanzania (pink). Panel (B) shows that *C. jejuni* transmission rates (orange) were always higher for children under 5 while *Shigella* transmission rates (light blue) were always higher for those aged 5 or above. Panel (C) shows that the community-to-household transmission rate (yellow) was always higher than the household transmission rate (dark blue) except for those aged 5 or older in Tanzania. Finally, panel (D) shows that children under 5 (circles) always had a higher transmission rate than those aged 5 or above (triangles) except for the transmission rate parameters corresponding to within-household *Shigella* transmission, which was only marginally higher for those aged 5 or above in Tanzania.

**7. Sensitivity analysis with a Ct cutoff of 35**

**Table S4:** Proportion of stool samples that had a cycle threshold of ≤35 and ≤ 27

| | | Bangladesh | | | Tanzania | | |
|---|---|---|---|---|---|---|---|
| | | Household members ≥5 | Children <5 | Total | Household members ≥5 | Children <5 | Total |
| % stools with Ct ≤ 35 | *Shigella* | 36.7 | 23.8 | 31.2 | 8.5 | 11.4 | 9.4 |
| | *C. jejuni* | 21.8 | 41.6 | 31.8 | 11.9 | 28.4 | 16.9 |
| % stools with Ct ≤ 27 | *Shigella* | 10.5 | 11.4 | 11.1 | 3.0 | 7.1 | 4.1 |
| | *C. jejuni* | 9.0 | 27.4 | 18.0 | 5.4 | 17.6 | 8.8 |

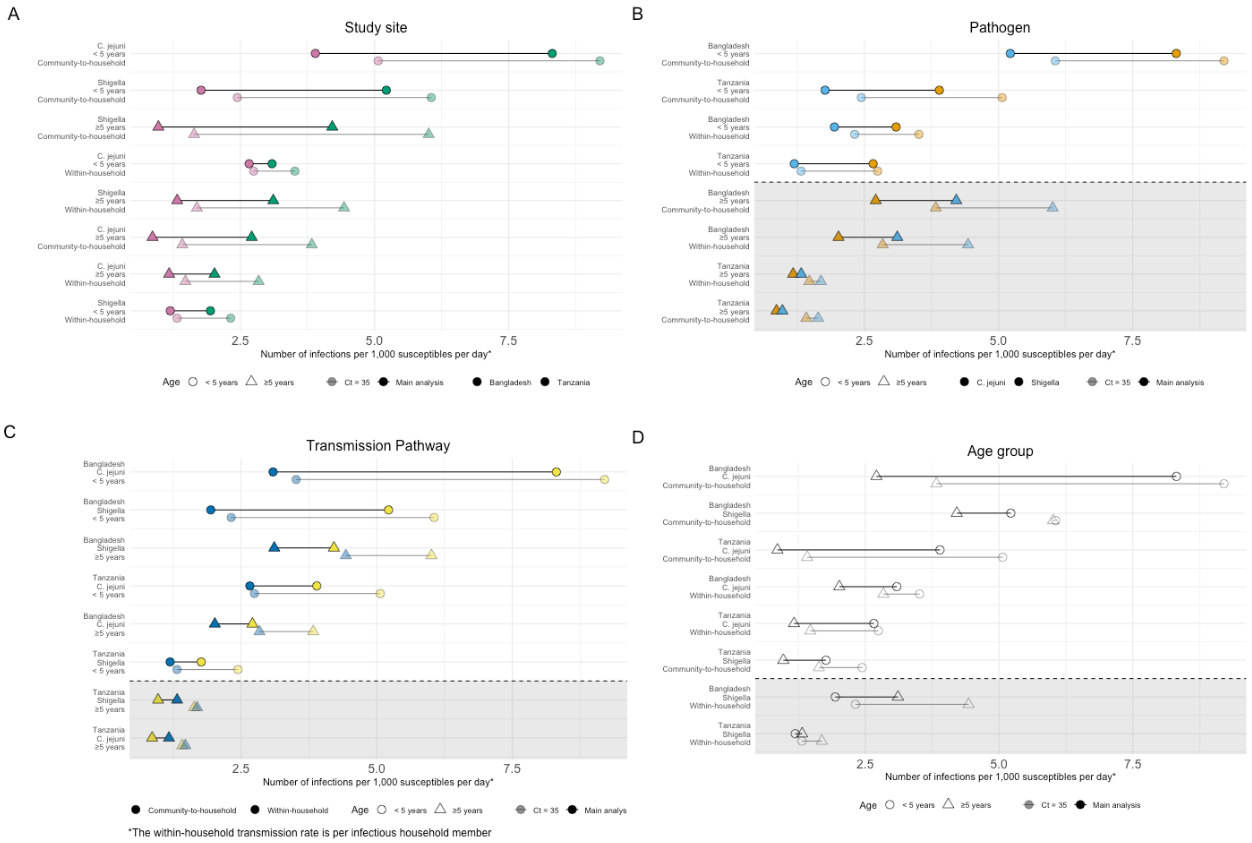


**Figure S6:** Sensitivity analysis showing the results from the main analysis (opaque) compared to the sensitivity analysis using a cycle threshold cutoff of 35 (translucent). Lollipop plots showing the median posterior value for each transmission rate parameter compared across pairs matched by study site (A), pathogen (B), transmission pathway (C), and age group (D). Each panel shows the same 16 transmission rate parameters rearranged according to the covariate of interest. Panel (A) shows that Bangladesh (green) always has a higher transmission rate than Tanzania (pink). Panel (B) shows that *C. jejuni* transmission rates (orange) were always higher for children under 5 while *Shigella* transmission rates (light blue) were always higher for those aged 5 or above. Panel (C) shows that the community-to-household transmission rate (yellow) was always higher than the household transmission rate (dark blue) except for those aged 5 or older in Tanzania. Finally, panel (D) shows that children under 5 (circles) always had a higher transmission rate than those aged 5 or above (triangles) except for the transmission rate parameters corresponding to within-household *Shigella* transmission, which was only marginally higher for those aged 5 or above in Tanzania.

#### 8. Sensitivity analysis with a Ct cutoff of 27

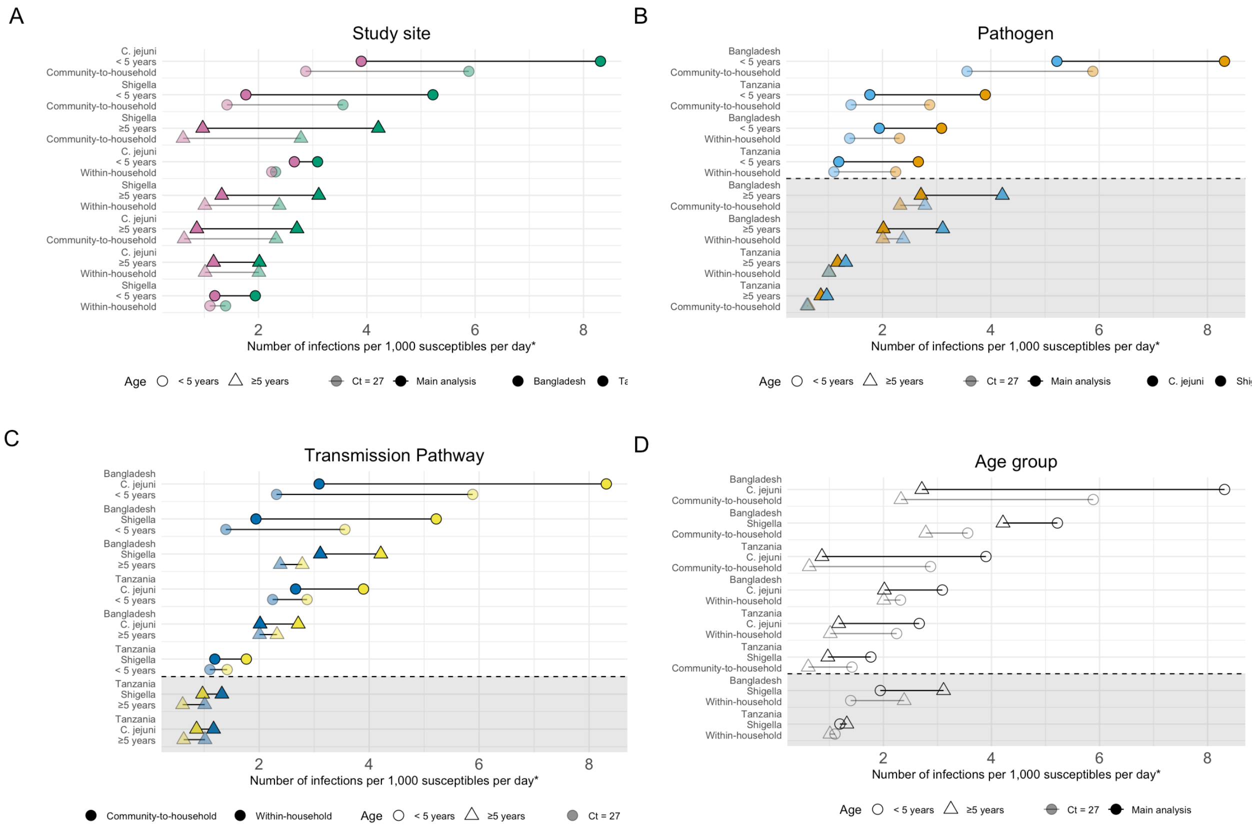


*The within-household transmission rate is per infectious household member

**Figure S7:** Sensitivity analysis showing the results from the main analysis (opaque) compared to the sensitivity analysis using a cycle threshold cutoff of 27 (translucent). Lollipop plots showing the median posterior value for each transmission rate parameter compared across pairs matched by study site (A), pathogen (B), transmission pathway (C), and age group (D). Each panel shows the same 16 transmission rate parameters rearranged according to the covariate of interest. Panel (A) shows that Bangladesh (green) always has a higher transmission rate than Tanzania (pink). Panel (B) shows that *C. jejuni* transmission rates (orange) were always higher for children under 5 while *Shigella* transmission rates (light blue) were always higher for those aged 5 or above. Panel (C) shows that the community-to-household transmission rate (yellow) was always higher than the household transmission rate (dark blue) except for those aged 5 or older in Tanzania. Finally, panel (D) shows that children under 5 (circles) always had a higher transmission rate than those aged 5 or above (triangles) except for the transmission rate parameters corresponding to within-household *Shigella* transmission, which was only marginally higher for those aged 5 or above in Tanzania.

**9. Sensitivity analysis of infection duration assumptions**

In the primary analysis, individuals with consecutive positive PCR tests at the monthly testing visits were assumed to remain continuously infectious between tests. To assess the sensitivity of our findings to this assumption, we generated alternative daily infection trajectories by assigning an infection episode to each positive PCR test. For each positive test, an infection duration was sampled from a distribution (see table below) and the infection start date was selected such that the observed positive test occurred within the sampled infection interval. Overlapping infection intervals were merged, while non-overlapping intervals were treated as distinct infection episodes. We considered exponential and gamma distributions with mean shedding durations of 20, 30, and 40 days applied equally to both pathogens. To additionally assess sensitivity to pathogen-specific differences in shedding duration, we considered four gamma-distributed scenarios where the mean shedding duration differed between the two pathogens.

| **Description** | **Distribution** | **Mean *Shigella* duration** | **Mean *C .jejuni* duration** | **Gamma shape** |
|---|---|---|---|---|
| Main analysis | Empirical | — | — | — |
| Short mean duration | Exponential | 20 | 20 | — |
| Intermediate mean duration | Exponential | 30 | 30 | — |
| Long mean duration | Exponential | 40 | 40 | — |
| Short mean duration | Gamma | 20 | 20 | 4 |
| Intermediate mean duration | Gamma | 30 | 30 | 4 |
| Long mean duration | Gamma | 40 | 40 | 4 |
| *Shigella* duration shorter | Gamma | 20 | 30 | 4 |
| *Shigella* duration much shorter | Gamma | 20 | 40 | 4 |
| *Shigella* duration longer | Gamma | 30 | 20 | 4 |
| *Shigella* duration much longer | Gamma | 40 | 20 | 4 |

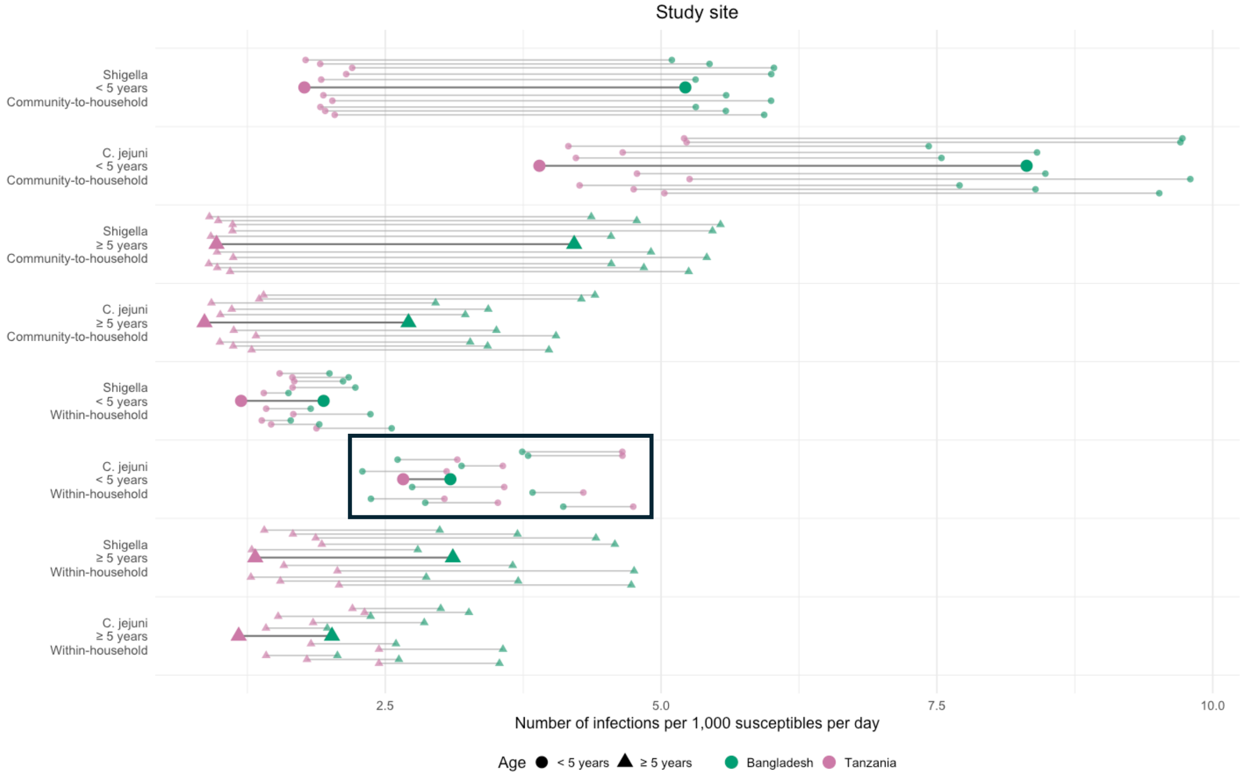


**Figure S8:** Results from the sensitivity analyses conducted testing the assumption of the infection duration with the alternative distributions described in the table above. Estimates from the main model from Figure 3A are shown in a larger size while each of the ten tested alternative infection length distributions are shown with the smaller shape sizes. Boxes are around pairs of transmission rate parameters where at least one alternative infection-duration assumption reversed the ordering of the paired estimates relative to the primary analysis.

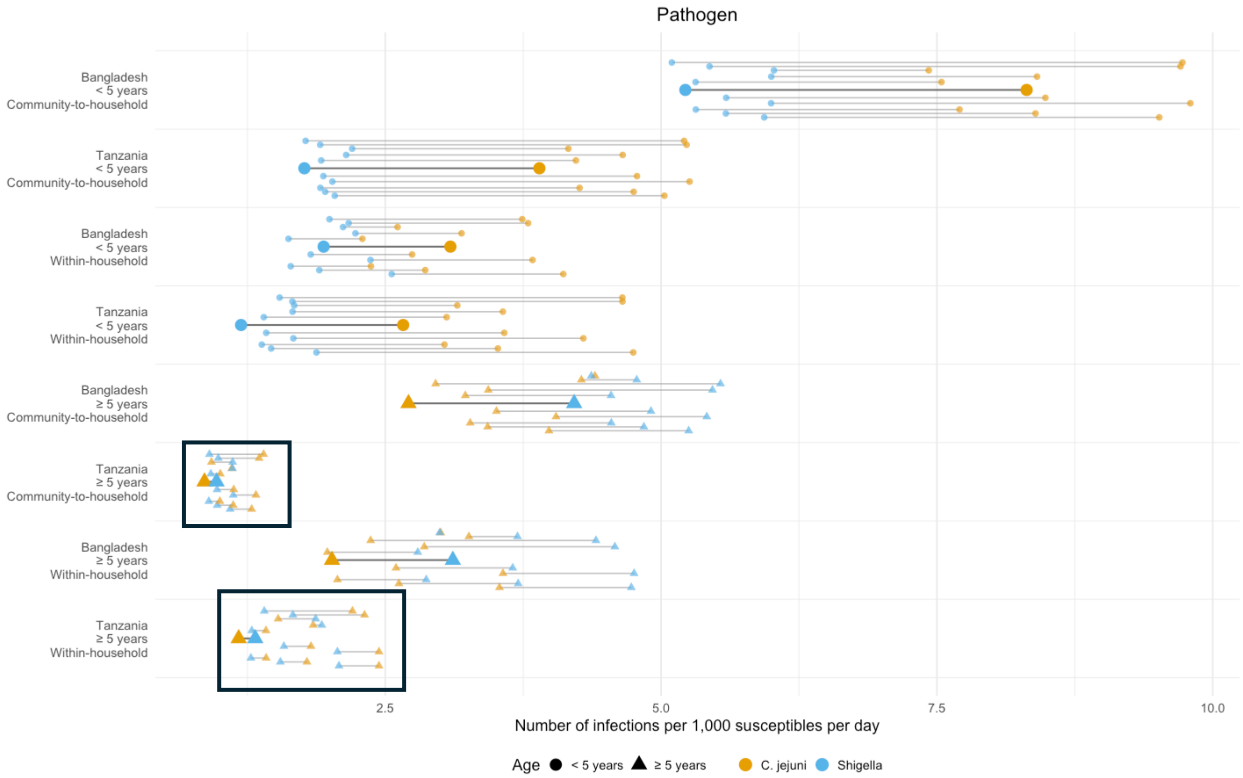


**Figure S9:** Results from the sensitivity analyses conducted testing the assumption of the infection duration with the alternative distributions described in the table above. Estimates from the main model from Figure 3B are shown in a larger size while each of the ten tested alternative infection length distributions are shown with the smaller shape sizes. Boxes are around pairs of transmission rate parameters where at least one alternative infection-duration assumption reversed the ordering of the paired estimates relative to the primary analysis.

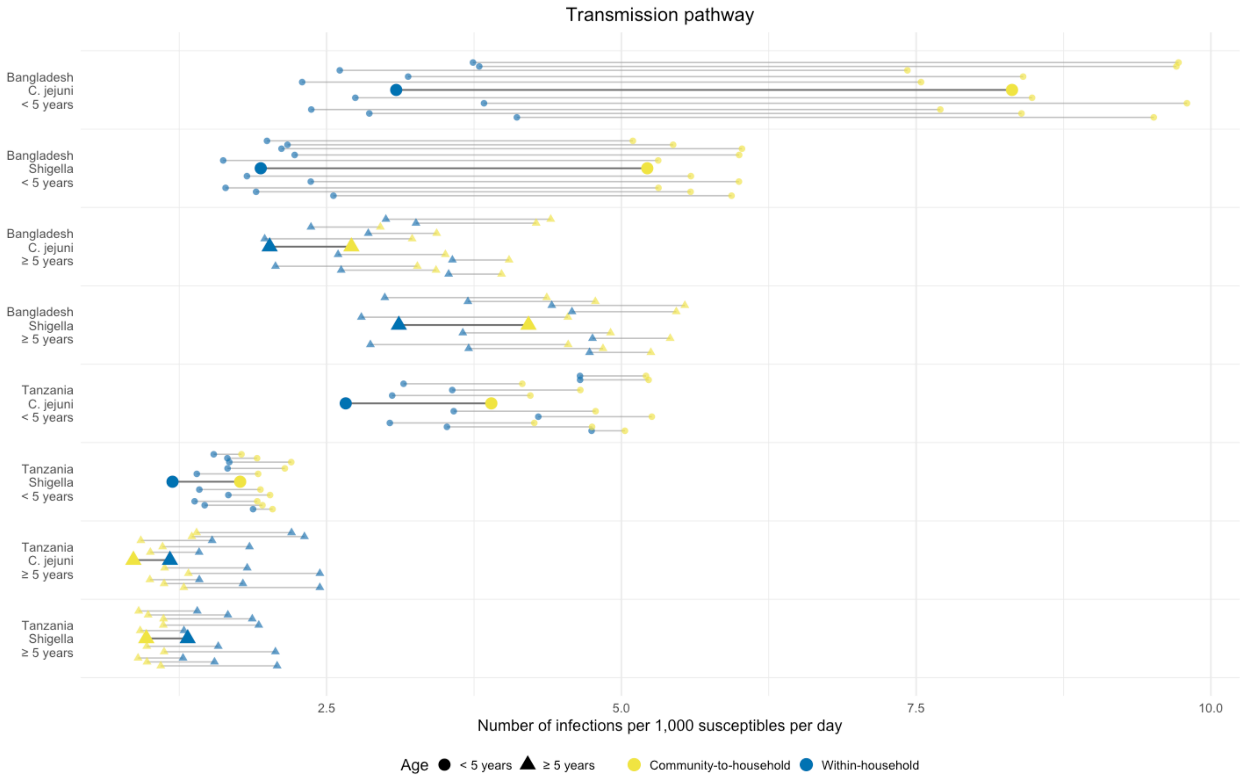


**Figure S10:** Results from the sensitivity analyses conducted testing the assumption of the infection duration with the alternative distributions described in the table above. Estimates from the main model from Figure 3C are shown in a larger size while each of the ten tested alternative infection length distributions are shown with the smaller shape sizes. Boxes are around pairs of transmission rate parameters where at least one alternative infection-duration assumption reversed the ordering of the paired estimates relative to the primary analysis.

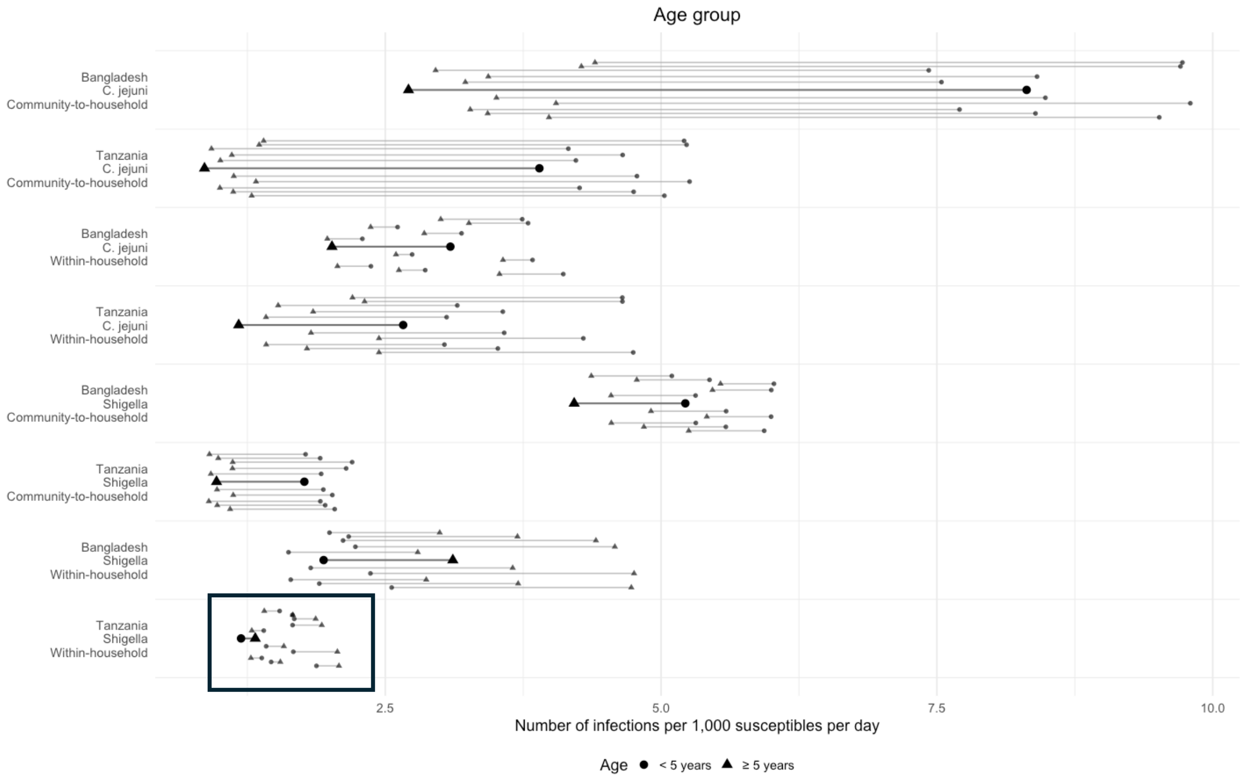


**Figure S11:** Results from the sensitivity analyses conducted testing the assumption of the infection duration with the alternative distributions described in the table above. Estimates from the main model from Figure 3D are shown in a larger size while each of the ten tested alternative infection length distributions are shown with the smaller shape sizes. Boxes are around pairs of transmission rate parameters where at least one alternative infection-duration assumption reversed the ordering of the paired estimates relative to the primary analysis.
